# Adverse drug withdrawal event signals in FAERS and Eudravigilance databases: a stratified disproportionality analysis study

**DOI:** 10.64898/2026.08.29.26361707

**Authors:** Zakir Khan, Caroline McCarthy, Kieran Dalton, Katharina Tabea Jungo, Ann Sinéad Doherty, Emily Reeve, Frank Moriarty

## Abstract

**Background:** Adverse drug withdrawal events (ADWEs) are a key safety concern during deprescribing but remain poorly explored in pharmacovigilance systems.

**Objectives:** To identify and compare ADWE signals across drug classes, different drugs within drug classes, and across patient characteristics, countries, and over time.

**Methods:** A case/non-case disproportionality analysis was conducted in FDA-FAERS and EMA-EudraVigilance pharmacovigilance databases, with stratification by age (adults: 18-64, older adults: ≥65), sex (male/female), reporting time (2004-2023 in 5-year intervals), and country (for EMA data). Disproportionality analysis (quantitative signal detection) was used to detect signals between ADWEs and drugs using the proportional reporting rate (PRR≥2), reporting odds ratio (ROR>1), and information component (IC>0) with case count ≥5.

**Results:** Overall, 158,501 reports (FDA-FAERS 145,514; EMA-EudraVigilance 12,987) included drug-event pairs related to ADWEs. In FDA-FAERS, clobetasone (IC=5.58; PRR=79.18; ROR=176.90) showed the strongest ADWE signals, followed by hydromorphone (4.85; 29.94; 37.37), hydrocodone, and paroxetine. In EMA-EudraVigilance, ethyl loflazepate (IC=6.01; PRR=119.80; ROR=197.53), clobetasone (5.39; 102.73; 155.10), veralipride, and levomethadone had the strongest signals. Most drugs maintained positive ADWE signals in analysis stratified into adults and older adults. However, among the top 10 drugs (based on highest IC values), buprenorphine/naloxone, desvenlafaxine, and baclofen in FDA-FAERS (ICs 4.95-6.05) showed stronger signals in older adults. A sex-based difference was observed, with paroxetine, venlafaxine, and buprenorphine/naloxone showing a stronger positive signal in females in both databases, whereas several opioids had stronger signals in males versus females across both databases.

**Conclusion:** This study suggests ADWE signals for some medications differ by age and sex, potentially indicating different risks for withdrawal effects.

## 1. Introduction

Adverse drug withdrawal events (ADWEs) are a subset of adverse drug reactions (ADRs) that are classified by the World Health Organisation’s adverse reaction terminology as type E reactions (withdrawal/end of use) and refer to signs and symptoms after the withdrawal of drugs [1, 2]. Fear of ADWEs is a key barrier to the implementation of deprescribing (the process of reducing or stopping unnecessary or harmful medications) in routine clinical practice [3, 4]. ADWEs can be categorised into physiological withdrawal reactions (often short-term and immediate) or the return of the condition being treated after drug discontinuation [3, 5, 6]. The severity of ADWEs can range from mild physical discomfort to severe psychological distress and even death. They differ by medicine type and may be influenced by treatment duration and patient characteristics [5, 7–9]. ADWEs have been reported after discontinuation of a wide range of drugs, including benzodiazepines, antidepressants, antipsychotics, narcotic analgesics, barbiturates, antihypertensives, corticosteroids, and proton pump inhibitors (5, 7, 21-24). However, evidence on these events to date is limited, as most studies have focused on single drug classes [10–12] and deprescribing trials rarely report ADWEs [3, 6].

Pharmacovigilance databases are an important source of drug safety information for ADR surveillance [13]. The analysis of individual case safety reports (ICSRs) in pharmacovigilance databases is essential for identifying previously unrecognised or unexpected ADRs in real-world clinical practice [14, 15]. Most countries designate a regulatory authority to monitor post-marketing drug surveillance. In the United States (US), the Food and Drug Administration (FDA) fulfils this role through the FDA Adverse Event Reporting System (FDA-FAERS) [16]. Similarly, the European Medicines Agency EudraVigilance (EMA-EudraVigilance) collects ADR reports in the European Economic Area (EEA) [17]. These databases are among the world’s largest collections of spontaneously reported ADRs, and their substantial amount of data, diversity, wide range of patient populations, and accessibility make them commonly used in pharmacovigilance research for the identification adverse event signals [18–21]. However, while these databases are routinely used for medication safety and ADRs caused by prescribing of medications [22], there appears to be limited use of this system for ADWEs caused by discontinuation or deprescribing [3, 5, 23–25].

Descriptive analyses of pharmacovigilance databases summarise reported frequencies without accounting for background reporting patterns and therefore cannot determine whether the frequency of report of an event with a drug reflects disproportional reporting; adverse events that are reported disproportionally more often for a particular medication, accounting for all of the events reported for that and other medications in the database. Therefore, descriptive findings alone may be insufficient and potentially misleading for the interpretation of drug-event relationships [26]. In contrast, the analysis of large pharmacovigilance datasets using disproportionality analysis enables the detection of clinically important safety signals. Disproportionality analysis is a quantitative method used for signal detection (to identify new, rare, or previously underrecognised risks associated with drugs) [27, 28]. Therefore, disproportionality analysis complements evidence obtained from pre-marketing clinical trials and observational studies, contributing to a more comprehensive assessment of drug safety [18, 20, 29]. A recent study evaluated ADWE signals associated with different drugs in the FDA-FAERS database [19], identifying that ADWEs were most often reported for individual drugs from opioid, antidepressant, and anti-anxiety drug classes. However, this disproportionality analysis study was limited to a database from one country and did not consider clinically informative grouping into drug classes. It also did not stratify analysis to examine differential signals across subgroups of patients.

Therefore, the aim of the current study was to detect signals of increased reporting of ADWEs when comparing drug classes and drugs within classes in US and European pharmacovigilance databases between 2004 and 2023. Additionally, we aimed to assess differential signals by age group (adult versus older adult), sex (male and female), country (for EMA-EudraVigilance data), and reporting trends over time (2004-2023) in 5-year intervals.

## 2. Methods

This study is reported according to the READUS-PV (The REporting of A Disproportionality analysis for drUg Safety signal detection using individual case safety reports in PharmacoVigilance) checklist [27]. The protocol for this study was preregistered on the Open Science Framework (https://doi.org/10.17605/OSF.IO/UTCWB) [30].

### 2.1. Study design

A case/non-case study of individual case safety reports using disproportionality analysis.

### 2.2. Databases

This study included data from two pharmacovigilance databases (FDA-FAERS and EMA-EudraVigilance). The FDA-FAERS, which was designed to support the FDA’s post-marketing safety assessment, includes data from ADRs, ADEs, and medication error reports. Quarterly data extract files (publicly available open data) were downloaded from the FDA’s website, covering the period from the first quarter of 2004 to the fourth quarter of 2023 [31]. Similarly, the EMA-EudraVigilance database is a repository of information on reports related to ADRs, ADEs, and medication error. Level 2A data (consists of anonymised detailed ICSRs) were obtained following approval from the EMA.

### 2.3. Cases and non-cases identification and key variables

Cases were all reports of relevant withdrawal events in all age groups. Events were identified using Preferred Terms (PTs) based on the Medical Dictionary for Regulatory Activities (MedDRA) [32–34]. We identified report records relating to ADWEs as those with a PT containing “withdraw”, and specific irrelevant terms were excluded (i.e. alcohol withdrawal syndrome, drug withdrawal syndrome neonatal, drug withdrawal maintenance therapy etc). Details of the different relevant PTs, non-relevant PTs, and key variables assessed in the FDA-FAERS and EMA-EudraVigilance databases are listed in ***Supplementary file 1 (Table S1.1 and S1.2)*.**

FDA categorises drugs as “primary” or “secondary” suspect, whereas EMA-EudraVigilance classifies drugs as “suspect” without distinguishing levels of attribution. We included all FDA-reported drugs in either category, aligning with the EMA [34, 35]. This approach reduces potential underestimation of cases and has been applied in previous FDA-based studies [36, 37]. Non-cases were reports of other non-withdrawal events (other ADRs and non-relevant withdrawal PTs).

### 2.4. Deduplication, drug cleaning, and mapping

We performed deduplication (removing duplicate entries for the same report, retaining the most recent record), as described in ***Supplementary file 1 (Table S1.3)***. Drugs in the FDA-FAERS files are identified by drug name (a mixture of entries, including valid trade names and verbatim text as reported, which contain pharmaceutical company names, dosage forms, or other irrelevant text) and prod_ai (product active ingredient). As prod_ai uniquely identifies drugs and was only introduced in the 2014Q3 extract [20], we extensively cleaned drugname for earlier years (2004Q1–2014Q2), removing irrelevant terms and mapping brand names to their generic equivalents, to derive prod_ai entries. The final cleaned prod_ai variable was used for analysis. We used RxNorm via RxNav (a browser for multiple drug information sources) and Drugs@FDA (the FDA-approved list of products for human use) to map drug brand names to their corresponding generic active ingredients [38, 39]. In the EMA-EudraVigilance, the drug name variable “RecodedProductsComps” (Active Substance, High Level) provided by EMA was used, which represents the standardised, non-proprietary name of the active product ingredients.

Drugs were classified according to the WHO Anatomical Therapeutic Chemical (ATC) system, using 5th-level ATC codes for individual drugs, and 3rd-level ATC codes for their respective drug classes. In cases of multiple ATC codes for a drug reflecting different indications, the code reflecting the most common clinical indication was used. ATC classification of individual drugs and drug classes in FDA-FAERS and EMA-EudraVigilance are described in ***Supplementary Excel files 1.1 and 1.2***, respectively.

### 2.5. Statistical analysis

We conducted analysis separately in both FDA-FAERS and EMA-EudraVigilance datasets. Descriptive analysis (frequency and percentage) was used to summarise demographic characteristics. Disproportionality analysis was conducted, computing the proportional reporting ratio (PRR), reporting odds ratio (ROR), and information component (IC), to identify safety signals between ADWEs and individual drugs [18–20]. A signal was considered significant if it met the criteria of ROR (>1), PRR (≥2), and IC (>0), indicating that a specific drug-ADWEs pair is reported more frequently than expected [18, 20, 40]. The thresholds applied to the estimates for PRR, ROR and IC are listed in ***Supplementary file 1 (Table S1.4)***. We set the minimum threshold for drug-event combination counts at ≥5 (rather than the previous standard of ≥3), which is recommended to enhance the efficiency and reliability of signal detection [26, 41]. We included all reports in the disproportionality analysis; however, only drug-ADWE combinations with ≥5 reports are presented and interpreted as signals.

Inter– and intra-drug class disproportionality analysis was conducted to assess differences in ADWE reporting signals between drug classes (by aggregating reports by class before analysis for those with ADWE reports) and between drugs within drug classes (by restricting analysis to drugs within each class with ADWE reports). To assess whether signals differed by demographics, geography or time, we also performed disproportionality analysis stratified by age groups (adult:18-64 years versus older adult: ≥65 years), sex (male and female), reporting trends over time data (grouped into 5-year intervals: 2004-2008, 2009-2013, 2014-2018, and 2019-2023), and country (for EMA-EudraVigilance data). Five-year intervals were selected to reduce the influence of year-to-year fluctuations in reporting patterns and allow more reliable comparisons of ADWE signals over broader time periods. Paediatric cases (0-17 years) and reports with missing age or sex information were included in the overall analyses but were not reported in stratified comparisons to ensure comparability between age and sex subgroups. Main results were presented for the drugs with the strongest signals in either subgroup (adults versus older adults or males versus females) and with signals in both subgroups. For the time-period analysis, results were presented for the drugs with the strongest ADWE signals in at least two time periods. Stata software (version 18) was utilised for data management, cleaning, and descriptive analysis. The pvda (Disproportionality Functions for Pharmacovigilance) package in R software was used for disproportionality analysis [42, 43]. All data management, cleaning and analysis scripts are available on **Zenodo** (https://zenodo.org/records/21710784).

## 3. Results

A total of 158,505 ADWE (FDA-FAERS: 145,514; EMA-EudraVigilance: 12,987) and 26,344,092 non-ADWE (FDA-FAERS: 20,587,882; EMA-EudraVigilance: 5,756,210) reports were identified. **Table 1** shows that a high proportion of reports had missing age data. Among reports with specified age, ADWEs in adults were more frequently reported than in older adults in both databases (FDA-FAERS=30.1% versus 5.8%; EMA-EudraVigilance=21.9% versus 4.7%). ADWEs reports were more commonly reported in females than males (FDA-FAERS=51.0% versus 41.2%; EMA-EudraVigilance=55.1% versus 42.4%). The highest proportion of ADWE reports were observed during 2019-2023 (FDA-FAERS=49.3%; EMA-EudraVigilance=38.3%).

**Table 1:** Details of ADWEs and Non-ADWEs reports in both databases.

| Variables | FDA-FAERS n (%) |  | EMA-EudraVigilance n (%) |  |
| --- | --- | --- | --- | --- |
|  | ADWEs cases<br>(n= 145,514) | Non-ADWEs<br>(n=20,587,882) | ADWEs cases<br>(n=12,987) | Non-ADWEs<br>(n=5,756,210) |
| **Age group** |  |  |  |  |
| Paediatrics | 3,114 (2.1) | 745,961 (3.6) | 269 (2.1) | 177,797 (3.1) |
| Adult | 43,833 (30.1) | 6,541,808 (31.8) | 2,842 (21.9) | 1,479,769 (25.7) |
| Older adults | 8,423 (5.8) | 4,122,482 (20.0) | 613 (4.7) | 494,892 (8.6) |
| Not Specified | 90,144 (61.9) | 9,177,631 (44.6) | 9,263 (71.3) | 3,602,752 (62.6) |
| **Sex** |  |  |  |  |
| Female | 74,163 (51.0) | 11,001,160 (53.4) | 7,153 (55.1) | 3,495,094 (60.7) |
| Male | 59,942 (41.2) | 7,202,470 (35) | 5,382 (42.4) | 2,054,129 (35.7) |
| Not Specified | 11,409 (7.8) | 2,384,251 (11.6) | 452 (3.5) | 206,987 (3.6) |
| **Time period** |  |  |  |  |
| 2004-2008 | 18,492 (12.7) | 1,716,071 (8.4) | 1,400 (10.8) | 374,076 (6.5) |
| 2009-2013 | 20,348 (14.0) | 3,566,818 (17.3) | 2,481 (19.1) | 719,879 (12.5) |
| 2014-2018 | 34,860 (24.0) | 6,543,590 (31.8) | 4,128 (31.8) | 1,318,790 (22.9) |
| 2019-2023 | 71,814 (49.3) | 8,761,403 (42.5) | 4,978 (38.3) | 3,343,465 (58.1) |

### 3.1. Disproportionality analysis based on individual drugs

A total of 144,218 reports (621 unique drugs after drug-event combination counts ≥5) from the FDA-FAERS and 16,343 reports (296 unique drugs) from the EMA-EudraVigilance were included in disproportionality analysis. In the FDA-FAERS database, clobetasone had the strongest signal (IC=5.58; PRR=79.18; ROR=176.9), followed by hydromorphone (4.85; 29.94; 37.37), hydrocodone, and paroxetine. In the EMA-EudraVigilance database, ethyl loflazepate (IC=6.01; PRR=119.80; ROR=197.53), clobetasone (5.39; 102.73; 155.10), veralipride, and levomethadone had the strongest signals (**Table 2**). Complete disproportionality metrics for all drugs are available for FDA-FAERS in ***Supplementary Excel files 2.1*** and for EMA-EudraVigilance in ***Supplementary Excel files 2.2***.

**Table 2:** Top 10 drugs with strong positive signals ranked based on IC value in FDA-FAERS and EMA-EudraVigilance databases.

| FDA-FAERS data |  |  |  |  |  | EMA-EudraVigilance data |  |  |  |  |  |
| --- | --- | --- | --- | --- | --- | --- | --- | --- | --- | --- | --- |
| Drug | ADWEs | Non-ADWEs | IC<br>(95% CI) | PRR<br>(95% CI) | ROR<br>(95% CI) | Drug | ADWEs | Non-ADWEs | IC<br>(95% CI) | PRR<br>(95% CI) | ROR<br>(95% CI) |
| Clobetasone | 60 | 48 | 5.58<br>(5.19-5.92) | 79.18<br>(66.88-93.73) | 176.9<br>(121.03-176.90) | Ethyl loflazepate | 70 | 107 | 6.01<br>(5.66-6.33) | 119.80<br>(99.79-143.83) | 197.53<br>(146.09-267.08) |
| Hydromorphone | 4226 | 16462 | 4.85<br>(4.81-4.90) | 29.94<br>(29.13-30.77) | 37.37<br>(36.11-38.67) | Clobetasone | 35 | 68 | 5.39<br>(4.88-5.83) | 102.73<br>(78.45-134.52) | 155.10<br>(103.13-233.24) |
| Hydrocodone | 3019 | 11800 | 4.85<br>(4.80-4.90) | 29.61<br>(28.67-30.58) | 36.93<br>(35.48-38.45) | Veralipride | 48 | 228 | 5.09<br>(4.66-5.47) | 52.61<br>(40.67-68.07) | 63.48<br>(46.48-86.69) |
| Paroxetine | 9585 | 39358 | 4.80<br>(4.77-4.82) | 29.79<br>(29.24-30.36) | 36.81<br>(35.97-37.66) | Levomethadone | 47 | 240 | 5.03<br>(4.59-5.41) | 49.54<br>(38.13-64.37) | 59.05<br>(43.18-80.75) |
| Oxycodone | 18824 | 81941 | 4.73<br>(4.71-4.75) | 30.41<br>(29.99-30.84) | 37.17<br>(36.55-37.80) | Buprenorphine and naloxone | 78 | 708 | 4.66<br>(4.32-4.96) | 30.07<br>(24.34-37.14) | 33.27<br>(26.32-42.06) |
| Betamethasone and fusidic acid | 17 | 12 | 4.63<br>(3.86-5.24) | 83.52<br>(61.51-113.4) | 200.43<br>(95.72-419.68) | Buprenorphine | 759 | 8777 | 4.56<br>(4.45-4.66) | 25.05<br>(23.36-26.87) | 27.13<br>(25.16-29.27) |
| Acetaminophen and dihydrocodeine | 22 | 40 | 4.58<br>(3.92-5.12) | 50.56<br>(36.14-70.72) | 77.81<br>(46.25-130.92) | Diamorphine | 181 | 2012 | 4.54<br>(4.33-4.75) | 25.15<br>(21.86-28.94) | 27.32<br>(23.45-31.83) |
| Duloxetine | 10274 | 51092 | 4.57<br>(4.54-4.60) | 25.58<br>(25.12-26.06) | 30.53<br>(29.87-31.20) | Methadone | 437 | 5300 | 4.48<br>(4.34-4.61) | 23.54<br>(21.49-25.79) | 25.40<br>(23.01-28.04) |
| Buprenorphine and naloxone | 3664 | 19074 | 4.51<br>(4.46-4.56) | 23.52<br>(22.82-24.24) | 27.85<br>(26.87-28.86) | Naloxone | 24 | 163 | 4.45<br>(3.82-4.97) | 38.77<br>(26.68-56.35) | 44.34<br>(28.87-68.07) |
| Morphine | 7656 | 43660 | 4.40<br>(4.37-4.43) | 22.37<br>(21.90-22.86) | 26.12<br>(25.48-26.78) | Hydromorphone | 77 | 932 | 4.33<br>(3.99-4.63) | 23.12<br>(18.64-28.67) | 24.95<br>(19.76-31.49) |

CI: Confidence interval; IC: Information component; PRR: Proportional reporting ratio; ROR: Reporting odds ratio

### 3.2. Inter-class disproportionality analysis

Disproportionality analysis between drug classes showed strongest signals in FDA-FAERS for corticosteroids combined with antibiotics (n=27; IC 4.83; PRR 57.91; ROR 127.76), opioids (n=57014; IC 3.57; PRR 19.01; ROR 21.31) and iron preparation (n=15; IC 2.74; PRR 18.54; ROR 22.30). The strongest signals in EMA-EudraVigilance were for antidotes (n=24; IC 4.08; PRR 25.56; ROR 29.17), drugs for constipation (n=22; IC 3.84; PRR 20.76; ROR 23.06) and psycholeptics-psychoanaleptics combinations (n=16; IC 3.54; PRR 17.50; ROR 19.09). Classes with strong signals across both databases included opioids (analgesics), muscle relaxants, anxiolytics, and drugs used in addictive disorder (including drugs used in opioid dependence such as buprenorphine/naloxone, methadone, levomethadone, and diamorphine, as well as other dependence-related medicines such as varenicline and nicotine) **(Figure 1).** Full results are reported in ***Supplementary Excel file 3.1 (***FDA-FAERS) and ***Supplementary Excel file 3.2*** (EMA-EudraVigilance).

**Figure 1:**
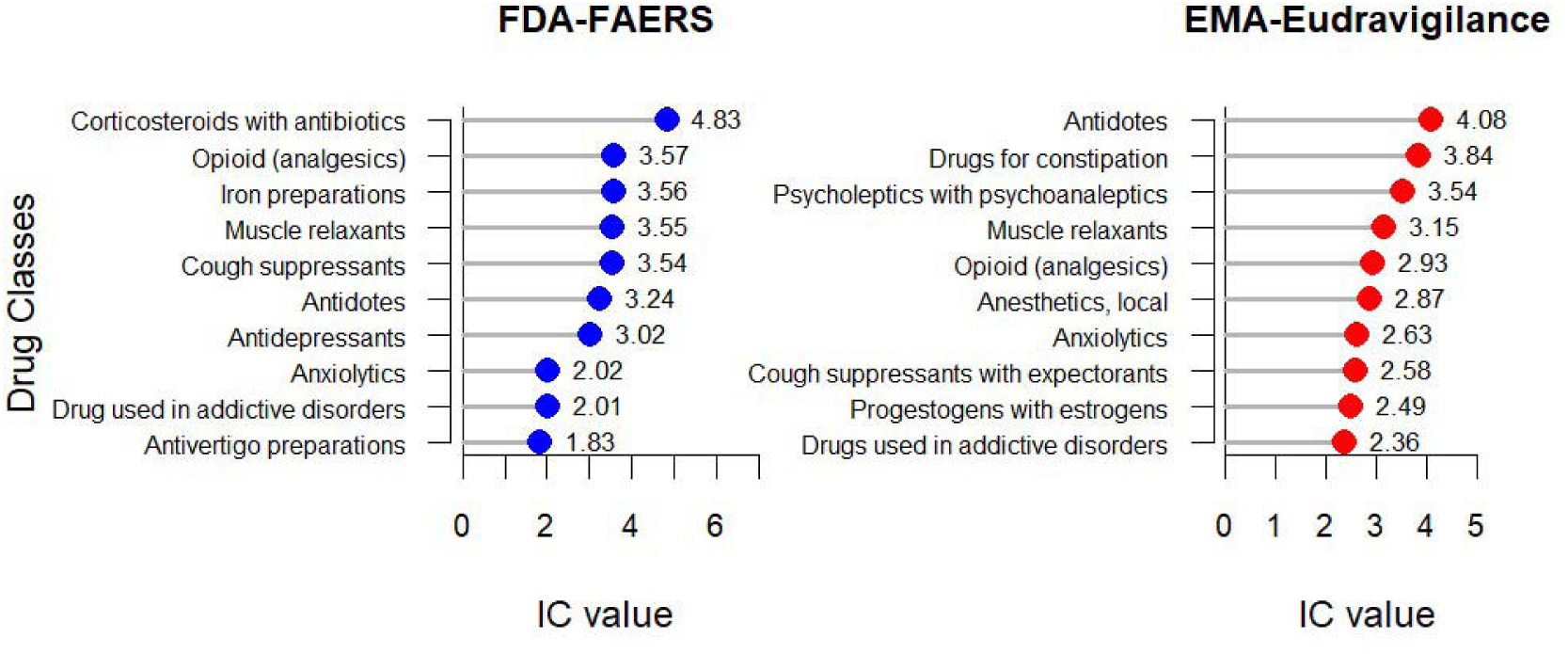
Inter-class disproportionality analysis ranked based on information criterion (IC) values in FDA-FAERS and EMA-EudraVigilance. ***Note:*** *Classes are defined using third-level ATC codes, i.e., “Opioids (analgesics)” refers to drugs included in N02A. Opioid-receptor modulators are also included in other third-level ATC groups (e.g., “Drugs used in addictive disorders” or “Cough suppressants”). Full details of drugs within each class are provided in Supplementary Excel Files 1.1, 1.2, 3.1 and 3.2*.

### 3.3. Intra-class disproportionality analysis

The intra-class disproportionality analysis identified differing signal strength for drugs relative to other agents within same therapeutic class, as well as between FDA-FAERS and EMA-EudraVigilance databases. Drugs were initially ranked based on IC values, and the ten drugs with the highest IC values were selected from each database. For each selected drug, the corresponding therapeutic class was identified, and the signal strength was compared with other agents within same therapeutic class. The top three drugs ranked by IC value within same therapeutic class are summarised in **Table 3**. For example, clobetasone showed the highest IC among corticosteroids in both databases (FDA-FAERS=5.86; EMA-EudraVigilance=5.05), whereas desonide and betamethasone demonstrated lower IC values in FDA-FAERS (IC=2.97 and 2.46, respectively), and clobetasol and hydrocortisone showed lower ICs in EMA-EudraVigilance (IC=2.12 and 1.48, respectively). The full intra-class disproportionality analysis results in FDA-FAERS and EMA-EudraVigilance are available in ***Supplementary Excel files 4.1 and 4.2***, respectively.

**Table 3:**
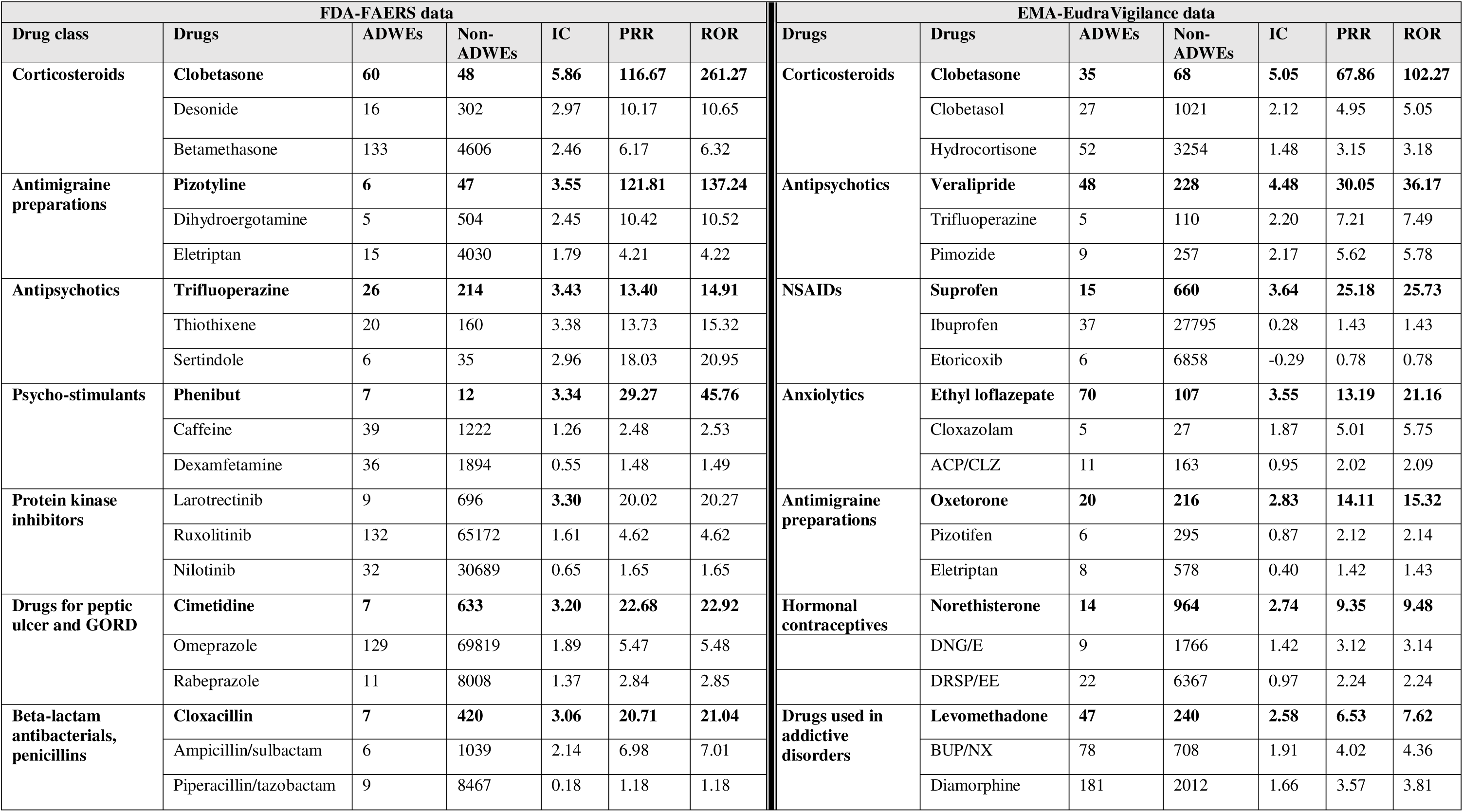
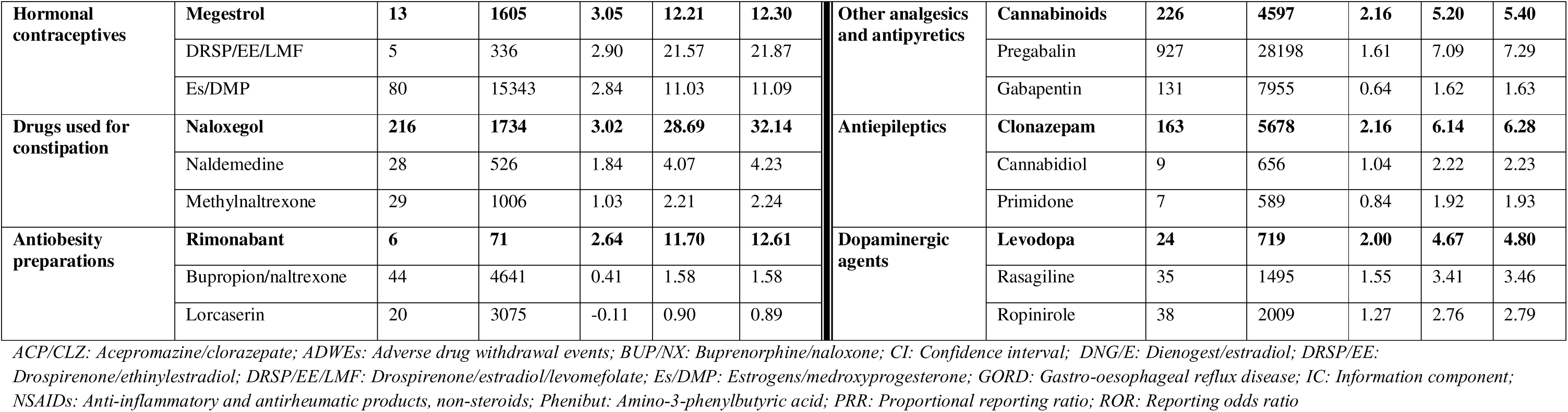
Intra-class disproportionality analysis ranked based on IC value in FDA-FAERS and EMA-EudraVigilance database.

### 3.4. Results by age group

Among the drugs with strongest signals, signal strength varied for ADWEs between adults (aged 18-64 years) and older adults (≥65 years). Buprenorphine/naloxone (IC=6.05), desvenlafaxine (IC=4.96), and baclofen (IC=4.95) in FDA-FAERS showed stronger signals in older adults. In contrast, paroxetine (IC=4.92) and naloxegol (IC=4.46) in FDA-FAERS and buprenorphine (IC=4.73), fentanyl (IC=4.16), and oxycodone (4.03) in EMA-EudraVigilance had stronger signals in adults **(Figure 2)**. Complete details are reported in the ***Supplementary Excel File 5.1*** *(FDA data)* and ***Supplementary Excel File 5.2*** *(EMA data)*.

**Figure 2:**
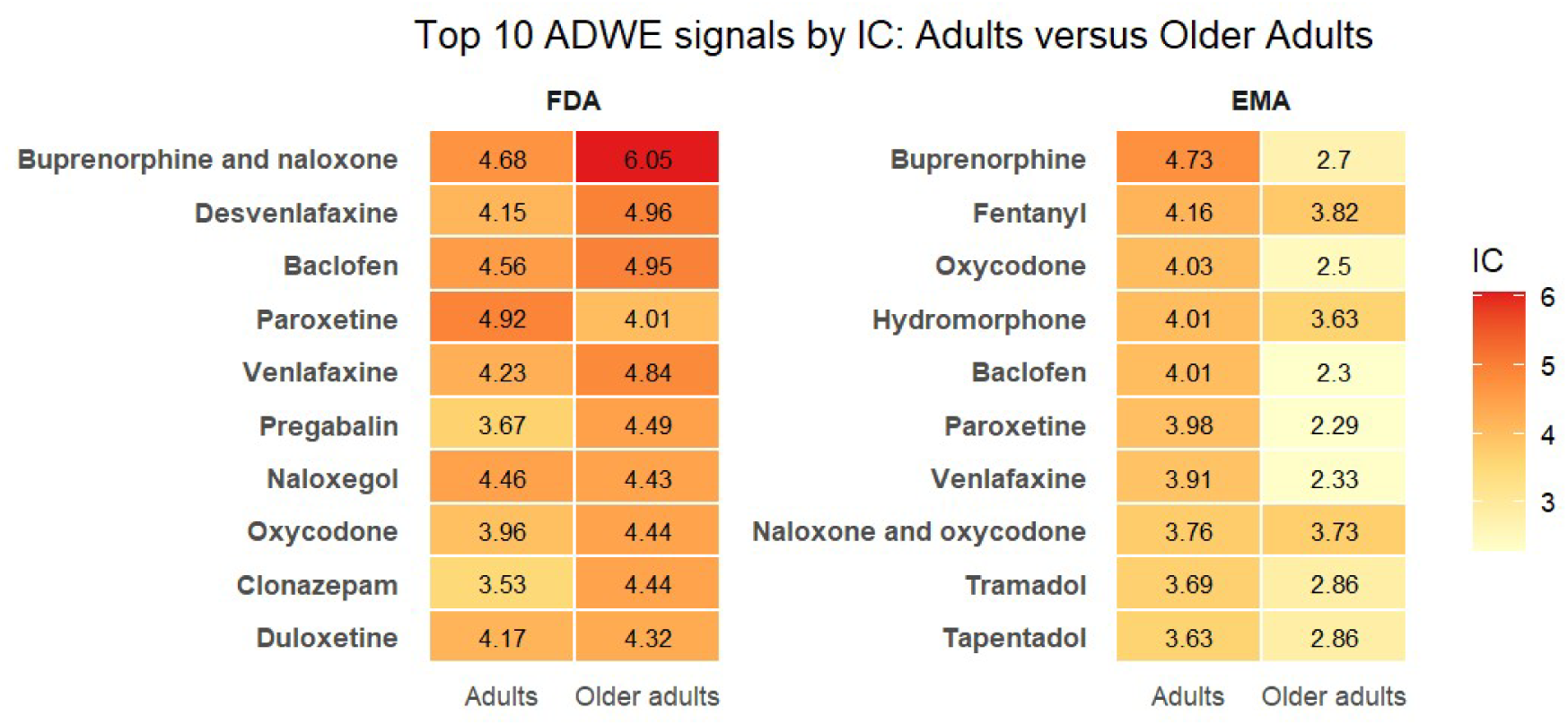
Information criterion (IC) values for drugs with the top 10 strongest signals by age group (adults 18-64 years and older adults ≥65 years) in FDA-FAERS and EMA-EudraVigilance database.

### 3.5. Results by sex

A sex-based difference was observed with stronger positive signals among females for paroxetine (IC=5.09), oxycodone (IC=4.83), duloxetine (IC=4.62), and baclofen (IC=4.32) in the FDA-FAERS database and ethyl loflazepate (IC=5.53), methadone (4.54), buprenorphine/naloxone (IC=4.36), and diamorphine (IC=4.28) in EMA-EudraVigilance database. A range of opioids had stronger signals in males versus females: hydromorphone (IC=5.11), hydrocodone (IC=5.03), and morphine (IC=4.56) in FDA-FAERS and buprenorphine (IC=4.33) and levomethadone (IC=4.32) in EMA-EudraVigilance **(Figure 3)**. Complete results by sex group are reported in ***Supplementary Excel File 6.1*** *(FDA data)* and ***Supplementary Excel File 6.2*** *(EMA data)*.

**Figure 3:**
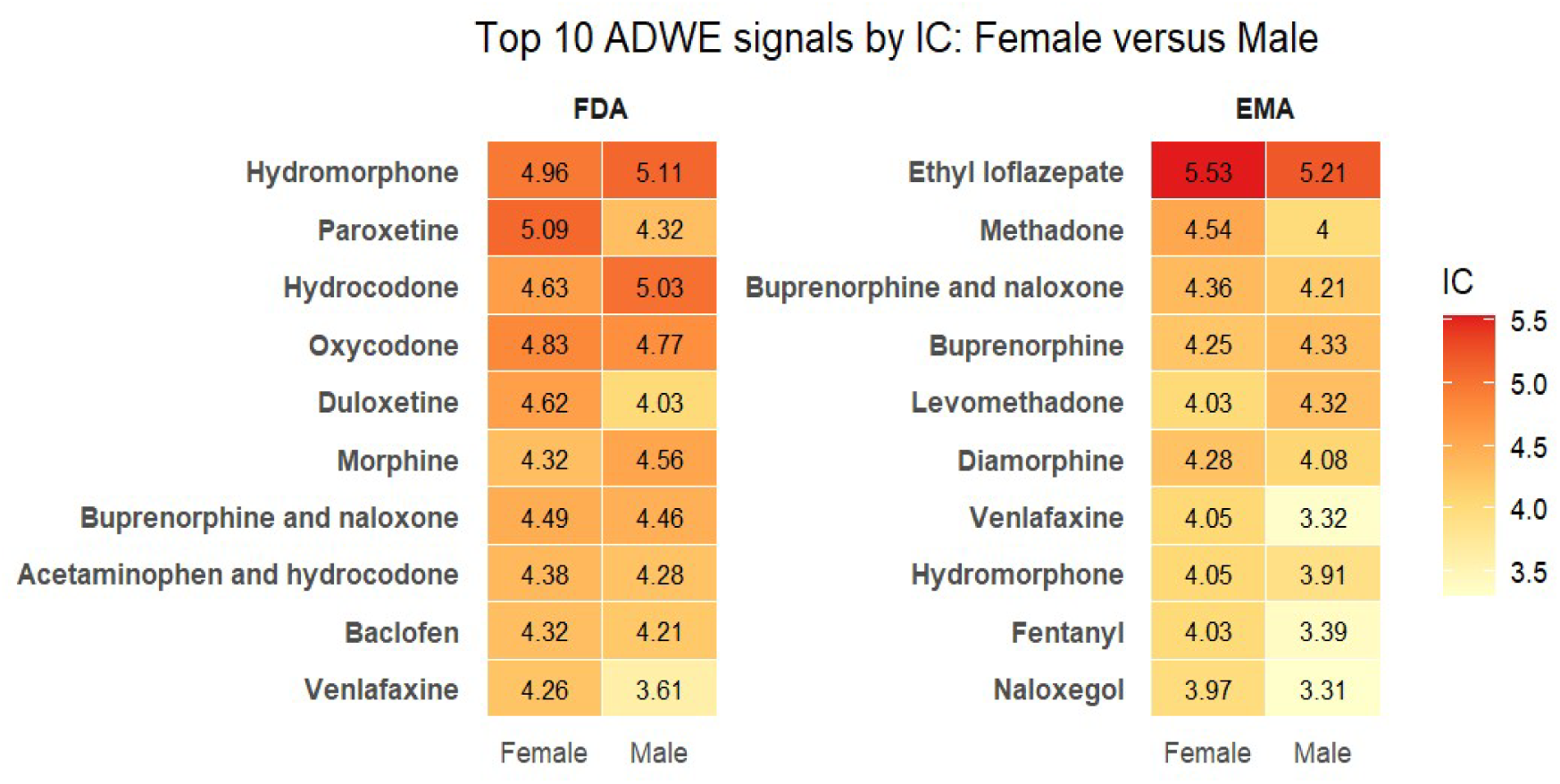
Information criterion (IC) values for drugs with the top 10 strongest signals by sex group in FDA-FAERS and EMA-EudraVigilance databases.

### 3.6. Results by time period

Analysis of the FDA-FAERS and EMA-EudraVigilance databases showed varied ADWE signals over time. Opioids (classified as analgesics or drugs for addictive disorders, FDA: hydrocodone, hydromorphone, oxycodone and morphine; EMA-EudraVigilance: levomethadone, buprenorphine, diamorphine and methadone) consistently represented the main drug class with strong signals. Among the top 10 signals over time, oxycodone, buprenorphine/naloxone, and baclofen consistently showed strong signals in FDA-FAERS database. Buprenorphine, methadone, and buprenorphine/naloxone consistently showed strong signals in EMA-EudraVigilance database. Some drugs, including clobetasone in both databases and diamorphine and estradiol/norethisterone in EMA-EudraVigilance showed strong signals in recent periods without signals in earlier periods **(Figure 4)**. Full details about all signals over time in FDA-FAERS and EMA-EudraVigilance are available in ***Supplementary Excel files 8.1*** and ***8.2***, respectively.

**Figure 4:**
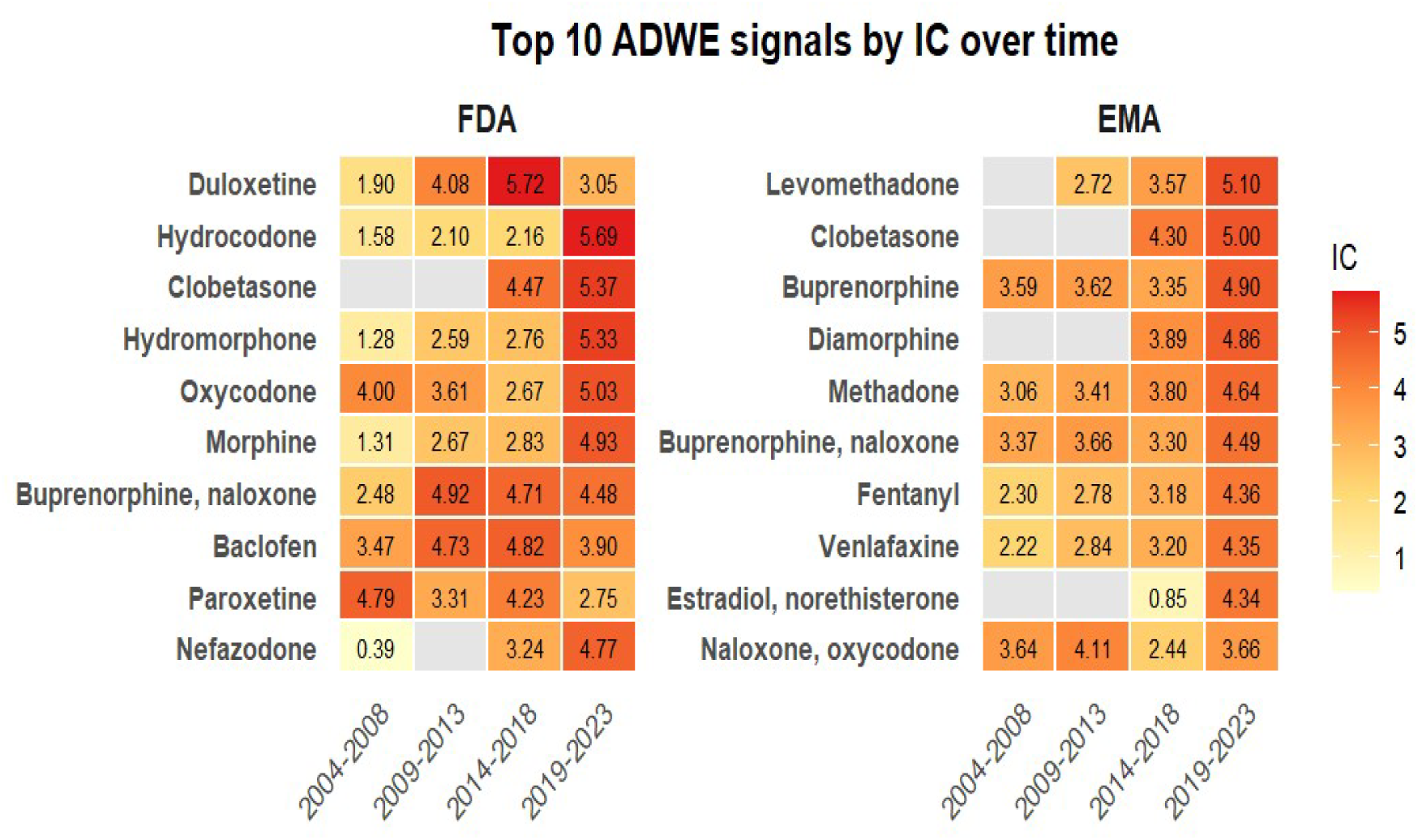
Information criterion (IC) values for drugs with the top 10 strongest signals over time in FDA-FAERS and EMA-EudraVigilance databases.

### 3.7. Results by country

In analysis across countries included in the EMA-EudraVigilance, the drugs with the strongest signals included opioids (levomethadone, naloxone/oxycodone, buprenorphine and methadone) and non-opioid drugs, including benzodiazepines/z-drugs (ethyl loflazepate, zolpidem, lorazepam), antidepressants (paroxetine, venlafaxine), antipsychotics (veralipride), baclofen, CNS-active drugs (rasagiline, levodopa), and corticosteroids (clobetasone). The highest IC observed was ethyl loflazepate in France (IC=5.34) followed by levomethadone in Germany (IC=4.87), rasagiline in Italy (IC=4.59), and clobetasone in UK (IC=4.40). The top three signals in the largest contributing countries are reported in **Table 4**, with full results included in ***Supplementary Excel File 7*.**

**Table 4:** Top three signals across countries in EMA-EudraVigilance data.

| Country | Drug | ADWEs | Non-ADWEs | IC | PRR | ROR |
| --- | --- | --- | --- | --- | --- | --- |
| France | **Ethyl loflazepate** | **64** | **78** | **5.34** | **59.67** | **107.81** |
|  | Buprenorphine | 385 | 2712 | 3.99 | 17.34 | 19.66 |
|  | Naltrexone | 21 | 125 | 3.73 | 18.90 | 21.91 |
| Germany | **Levomethadone** | **43** | **195** | **4.87** | **44.56** | **54.17** |
|  | Naloxone and oxycodone | 71 | 667 | 4.33 | 23.92 | 26.36 |
|  | Fentanyl | 195 | 2165 | 4.26 | 21.34 | 23.18 |
| Italy | **Rasagiline** | **29** | **136** | **4.59** | **42.27** | **51.07** |
|  | Levodopa | 14 | 100 | 3.85 | 28.73 | 32.62 |
|  | Methadone | 23 | 299 | 3.62 | 16.96 | 18.19 |
| UK | **Clobetasone** | **25** | **43** | **4.40** | **35.89** | **56.18** |
|  | Paroxetine | 117 | 420 | 4.28 | 21.92 | 27.74 |
|  | Naloxone and oxycodone | 24 | 63 | 4.13 | 26.92 | 36.79 |
| Spain | **Veralipride** | **45** | **182** | **4.19** | **24.83** | **30.72** |
|  | Buprenorphine | 33 | 345 | 3.13 | 10.61 | 11.53 |
|  | Baclofen | 13 | 147 | 2.82 | 9.50 | 10.25 |
| Poland | **Zolpidem** | **44** | **248** | **3.83** | **19.32** | **22.57** |
|  | Lorazepam | 14 | 83 | 3.40 | 16.83 | 19.50 |
|  | Chlorazepate | 6 | 40 | 2.83 | 14.85 | 16.93 |
| Netherlands | **Clobetasone** | **8** | **7** | **3.77** | **66.67** | **141.73** |
|  | Baclofen | 31 | 216 | 3.66 | 15.98 | 18.13 |
|  | Venlafaxine | 112 | 1340 | 3.20 | 10.48 | 11.28 |
| Sweden | **Paroxetine** | **16** | **154** | **2.76** | **8.47** | **9.25** |
|  | Venlafaxine | 62 | 822 | 2.57 | 6.68 | 7.11 |
|  | Tramadol | 36 | 608 | 2.22 | 5.14 | 5.39 |
| Finland | **Amitriptyline and chlordiazepoxide** | **6** | **13** | **2.63** | **11.27** | **16.01** |
|  | Paroxetine | 5 | 10 | 2.56 | 11.85 | 17.28 |
|  | Methadone | 6 | 15 | 2.56 | 10.19 | 13.87 |
| Denmark | **Tramadol** | **67** | **286** | **2.60** | **7.71** | **9.28** |
|  | Estradiol and norethisterone | 14 | 54 | 2.50 | 7.10 | 8.68 |
|  | Oxazepam | 7 | 57 | 1.62 | 3.69 | 4.02 |

IC: Information component; PRR; Proportional reporting ratio; ROR: Reporting odds ratio.

## 4. Discussion

To the best of our knowledge, no previous study has conducted an analysis of ADWEs across age groups, sex, and over time, or compared reporting between the FDA-FAERS and EMA-EudraVigilance databases. Therefore, this has filled an important evidence gap on ADWE reporting patterns and risks across different drugs. In this study, strong signals were identified for different centrally acting drugs across both databases. Opioid-related medications had consistently strong signals in both databases, including opioid analgesics classified under the ATC *Opioids (N02A)* category (e.g., hydromorphone, oxycodone, and morphine in FDA-FAERS), as well as opioid-related medicines classified under *Drugs used in addictive disorders (N07B)* for opioid dependence treatment (e.g., levomethadone, buprenorphine/naloxone, diamorphine, and methadone in EMA-EudraVigilance). A previous 10-year analysis of opioid-related ADWEs in the FDA-FAERS and EMA-EudraVigilance database identified tramadol and oxycodone as more strongly associated with drug withdrawal symptoms than other opioids [44]. The present findings reinforce the value of pharmacovigilance databases as important real-world data sources for monitoring opioid-related ADWEs and highlight the need for clinicians to remain aware of these events in routine practice.

In this study, strong ADWE signals were identified for other psychotropic drugs including antidepressants (paroxetine and duloxetine in FDA-FAERS), anxiolytics (ethyl loflazepate in EMA-EudraVigilance), and antipsychotics (veralipride in EMA-EudraVigilance). These findings are consistent with previous pharmacovigilance studies focused on antidepressant and selective serotonin reuptake inhibitor (SSRI) ADWE reports in global, regional, and national databases [10, 12, 45, 46], as well as research on antipsychotics in the WHO database [11]. As psychotropic drugs are well recognised to be associated with withdrawal reactions and may also be associated with relapse or return of condition [47–49], prescribers should therefore consider risk of ADWEs from psychotropic drugs throughout the treatment, including during initiation, continuation, and discontinuation. Strong signals were also observed for topical corticosteroids, particularly clobetasone across both databases. This finding is less commonly highlighted in prior pharmacovigilance studies [19, 50]. Previous evidence sources, including systematic review [52] and regulatory warning [53], have reported withdrawal reactions associated with topical corticosteroids, including clobetasone, beclometasone, and clobetasol. Similarly, clobetasone’s patient information leaflet recognises “*steroid withdrawal reaction*” as an adverse effect with an unknown frequency, indicating insufficient data for incidence estimation, although symptoms may occur within days to weeks after discontinuation [54]. The current study provides additional real-world evidence supporting this clinically recognised but poorly quantified adverse reaction. The strength of the clobetasone signal increased over time, with the IC rising from 4.47 (2014-2018) to 5.37 (2019-2023) in FDA-FAERS and from 4.30 to 5.00 in EMA-EudraVigilance. This increase may partly reflect growing awareness of topical corticosteroid withdrawal reactions, including discussions through online patient communities and social media, which may have influenced reporting behaviour [55, 56]. However, this finding should be interpreted cautiously, as spontaneous reporting data cannot determine whether the increased signal reflects greater reporting of recognised events or a true increase in clinical risk. Therefore, increased vigilance among clinicians and patients and clear communication regarding the need for gradual tapering after prolonged use of potent corticosteroids may support earlier recognition, reporting and appropriate management of corticosteroid-related withdrawal reactions.

This study also demonstrated that high reporting frequency did not necessarily correspond to strong disproportionality signals. Several drugs with comparatively lower reporting frequencies, including clobetasone, hydromorphone, betamethasone/fusidic acid, ethyl loflazepate showed strong signals. In contrast, our previous descriptive analysis of ADWE reporting patterns in the FDA-FAERS and EMA-EudraVigilance databases identified several frequently reported drug classes/drugs; however, many of the most commonly reported drug classes and drugs (e.g., oxycodone, buprenorphine, duloxetine, venlafaxine, pregabalin) did not consistently exhibit the strongest disproportionality signals [34]. These findings highlight the complementary value of descriptive and disproportionality analyses; descriptive analysis provides information on the frequency and characteristics of ADWE reports, while disproportionality analysis identifies potential signals that may not be apparent from reporting frequency alone.

Stratified analysis by age showed higher signals among older adults for drugs such as buprenorphine/naloxone, desvenlafaxine, baclofen, venlafaxine, pregabalin, oxycodone, and clonazepam in the FDA-FAERS, which may reflect age-related pharmacokinetic/pharmacodynamic changes, higher drug exposure, and polypharmacy [57–59]. However, despite these stronger signals for individual drugs, the overall number of reported withdrawal reactions was lower among older adults than adults [34]. Whether these stronger signals reflect greater susceptibility to withdrawal reactions among older adults is unclear, as previous evidence specifically examining age-related differences in ADWE signals is limited. The observed differences may also reflect greater multimorbidity and treatment complexity in older adults, as well as difficulties distinguishing withdrawal reactions from return of condition. Secondly, this study’s sex-stratified analyses revealed stronger ADWE signals for antidepressants (e.g., paroxetine, duloxetine, and venlafaxine) in females than in males, potentially reflecting sex-related differences in pharmacokinetics, as well as both sex and gender-related differences in the prevalence of depression and anxiety disorders and antidepressant prescribing. Social norms and gender roles, including greater caregiving responsibilities among women and lower treatment-seeking behaviour among men, may also contribute to these differences.[60–63] In contrast, opioid-related ADWE signals showed mixed sex-specific patterns across drugs in both databases. Hydromorphone, hydrocodone, morphine, and levomethadone showed relatively stronger signals in males, whereas methadone, fentanyl, and diamorphine demonstrated stronger signals in females. Sex differences in opioid-related ADRs have previously been reported with stronger signals in females for some opioids, including codeine and fentanyl [64]. Overall, the patterns of signals observed in stratified analyses were not always consistent across both databases, which may reflect differences in prescribing practices, reporting behaviour, exposure distribution, and healthcare systems rather than differences in biological responses between the two regions. Moreover, to our knowledge, no previous studies have specifically examined ADWE signals using age– and sex-stratified disproportionality analyses. Previous pharmacovigilance studies have employed age-stratified analyses to examine ADRs related to pain therapies and clarithromycin [40, 65], and medication errors [66]. These findings highlight the importance of stratified analysis of pharmacovigilance data based on patient characteristics to enhance signal interpretation and identify potentially susceptible patient populations.

Our analysis stratified by time period showed that several drug classes consistently demonstrated strong signals across databases. Opioid-related medications showed strong signals, which may reflect the growing prevalence of use globally and across the US and Europe, more awareness and efforts to address the opioid epidemic [67, 68]. In FDA-FAERS, signals remained relatively consistent over time and were primarily driven by prescription opioid analgesics, alongside CNS-active drugs such as baclofen and venlafaxine. In contrast, EMA-EudraVigilance showed more marked changes in signal composition over time, with increasing prominence of opioid substitution therapies, particularly buprenorphine, methadone, and diamorphine in recent periods, as well as signals for benzodiazepine-related (e.g., ethyl loflazepate) and topical agents (e.g., clobetasone). These temporal differences may reflect changes in drug utilisation, and practices around stopping medicines. Although increased utilisation may increase the number of withdrawal reports, it would not necessarily result in stronger disproportionality signals because these measures assess relative reporting rather than the absolute number of reports. More frequent discontinuation could increase occurrence and reporting of withdrawal reactions, while changes towards safer deprescribing practices, including appropriate tapering and monitoring, could reduce their occurrence. Additionally, increased cessation without appropriate support could potentially increase withdrawal reactions. Increased awareness and recognition of ADWEs may also contribute to changes in reporting over time. However, the relative contribution of these factors cannot be distinguished using spontaneous reporting data. Some of our findings indicate potential signals for drugs not widely implicated in withdrawal reactions previously, such as ethyl loflazepate, veralipride, and baclofen. Ethyl loflazepate demonstrated a strong disproportionality signal with prominence in France during 2019-2023, consistent with known withdrawal risk but rarely reported in pharmacovigilance literature [69, 70]. Dopaminergic agents (rasagiline and levodopa) also showed consistent signals, however with limited literature and regulatory warnings [71–74].

These extend beyond CNS drugs, with suprofen, larotrectinib, cimetidine, piroxicam, and cetirizine showing weaker but potentially meaningful signals (see Supplementary Excel files 2.1 and 2.2), warranting further investigation. Overall, these findings indicate that clinically important ADWEs may occur even among drugs with relatively low reporting frequencies. Therefore, clinicians should consider the risk of withdrawal reactions in any patients taking prescription or over-the-counter medications.

### 4.1. Strengths and Limitations

This study uses two large regional pharmacovigilance databases to provide novel evidence on ADWE signals for medications, which complements the limited evidence on these events from deprescribing trials and other studies. It is also the first study to consider age– and sex-stratified signals for ADWEs. This study provides detailed methodological documentation from data retrieval to disproportionality analysis, including analytical code for both databases (available as supplementary materials), enabling researchers to reproduce, validate, and extend the findings of this work.

However, some limitations should be considered in interpreting results, as reported events do not establish causality, and may be influenced by the underlying disease, concomitant medications, and incomplete information on treatment duration and discontinuation. Moreover, spontaneous reporting systems are subject to under-reporting, reporting bias, and variability in data completeness and quality. Signals derived from these databases should be interpreted as hypothesis-generating rather than evidence of a causal relationship and require confirmation using complementary evidence such as epidemiological studies. Disproportionality measures only assess a risk of reporting and are not representative of the true risk, and signals may be influenced by prescribing trends, media attention, regulatory warnings, or differences in pharmacovigilance practices across regions and time periods. Additionally, reporting bias may be particularly relevant for drugs perceived as having a low ADR risk (e.g., topical corticosteroids), where fewer overall reports may result in a smaller denominator for disproportionality analyses, thus amplifying signals for ADWEs. Increased awareness of topical corticosteroid withdrawal through social media may have influenced reporting behaviour and the observed strength of ADWE signals [55]. The use of the ATC classification system also presents limitations when interpreting drug-class findings. Although the ATC system provides a standardised and reproducible framework for drug classification, medicines with similar pharmacological properties may be assigned to different therapeutic classes according to their indication or formulation. For example, some drugs with opioid pharmacological activity are classified outside N02A (analgesics), such as drugs used in addictive disorders or cough suppressants. Similarly, other classifications may include drugs whose pharmacological or clinical use may not be immediately apparent from the class label, such as cocaine and ketamine being classified as local and general anaesthetics, respectively. These classification characteristics should therefore be considered when interpreting drug-class signals, particularly where a class is predominantly driven by one or a small number of medicines. Stronger signals observed in older adults may partly reflect higher medication exposure, polypharmacy, or increased healthcare contact rather than biological susceptibility alone. Only reports with complete demographic information were included in stratified analyses, while paediatric reports and reports with missing age or sex were excluded, which may have affected subgroup representativeness and generalisability of the findings. However, disproportionality analysis-based studies play a crucial role in drug safety, and can inform risk management strategies, ongoing post-market surveillance, comparative safety analysis, and clinical decision-making to ensure patient safety and optimal therapeutic outcomes. Finally, spontaneous reporting data cannot reliably distinguish withdrawal reactions from return of condition after discontinuation of a drug. The identified ADWE signals should be interpreted cautiously, as they may reflect withdrawal reactions, return of condition, or both.

### 4.2. Clinical Implications and Future Research

The findings have important implications for psychotropic drug use. The strong signals identified for antidepressants, benzodiazepines, antipsychotics, and opioids are clinically important given the common and sometimes potentially inappropriate long-term use of these drugs in routine practice, including antipsychotic use for behavioural and psychological symptoms of dementia and widespread SSRI prescribing [75–77]. Risk of withdrawal reactions and other adverse effects should be considered and weighed against sometimes marginal benefits of these medications when considering treatment initiation and should be part of the informed consent and shared-decision making processes. These findings reinforce the importance of careful monitoring, gradual tapering strategies, and patient-centred deprescribing approaches and clinician awareness to minimise withdrawal-related harms. The observed sex– and age-related differences further suggest that both prescribing and deprescribing decisions may need to be individualised, taking into account patient susceptibility and treatment exposure patterns. The detection of less expected signals in non-psychotropic drugs – including clobetasone (topical corticosteroid), suprofen and piroxicam (NSAIDs), and cetirizine (antihistamine) – reinforces that withdrawal reactions are not limited to centrally-acting drugs. This expands the clinical relevance of ADWEs beyond psychotropic prescribing and indicates that clinicians should remain alert to withdrawal phenomena, even with commonly used topical or over-the-counter therapies. The identification of topical corticosteroids as potential ADWE signals highlights the clinical importance of recognising withdrawal reactions beyond psychotropic medications. For topical corticosteroids, withdrawal symptoms can occur rapidly and be distressing, whereas other adverse effects, such as skin thinning or pigmentation changes, typically develop gradually. If clinicians and patients do not recognise withdrawal reactions, withdrawal signs/symptoms may be mistaken for recurrence of the underlying condition, potentially leading to unnecessary reinitiation of treatment and failure of deprescribing efforts. Future research should focus on validating these signals in longitudinal cohort studies, and prospective deprescribing trials. Additionally, further studies are also needed to better understand withdrawal mechanisms across different drugs, and to explore patient-level predictors of susceptibility, including dose, duration of use, and comorbidity profiles. Improved standardisation of ADWE reporting in pharmacovigilance systems would also enhance signal detection and comparability across subgroup analysis [78]. Moreover, these findings suggest that prescribing and deprescribing guidelines should incorporate consideration of potential ADWEs associated with individual medicines and patient characteristics when informing decisions about treatment initiation, continuation and discontinuation.

## 5. Conclusion

This study suggests ADWE signals for some medications may differ by age and sex, potentially indicating different risks of withdrawal effects in different demographic groups. This study provides supportive evidence of known ADWEs with some drug classes, and also identifies some novel signals not previously reported to help inform appropriate deprescribing strategies. Further confirmation of the ADWE signals identified in this study is needed using epidemiological and clinical evidence. More research to clarify underlying mechanisms and develop effective strategies to prevent and mitigate ADWEs would support efforts to reduce medication harm.

## Abbreviations

*ADRs: Adverse drug reactions; ADEs: Adverse drug events; ADWEs: Adverse drug withdrawal events; ATC: Anatomical Therapeutic Chemical; CI: Confidence interval; CNS: Central nervous system; EMA: European Medicines Agency; EEA: European Economic Area; FAERS: FDA Adverse Event Reporting System; FDA: Food and Drug Administration; GORD: Gastro-oesophageal reflux disease; IC: Information component; ICSRs: Individual case safety reports; MedDRA: Medical Dictionary for Regulatory Activities; NSAIDs: Non-steroidal anti-inflammatory drugs; PRR: Proportional reporting ratio; PTs: Preferred Terms; READUS-PV: The REporting of A Disproportionality analysis for drUg Safety signal detection using individual case safety reports in PharmacoVigilance; ROR: Reporting odds ratio; SSRI: Selective serotonin reuptake inhibitor; US: United States; WHO: World Health Organization*.

## Declarations

### Funding and disclaimer

This research is funded by the European Union through the Marie Skłodowska-Curie Actions (MSCA) programme (Project acronym: HEAD-P; Project number: 101149577) https://cordis.europa.eu/project/id/101149577. Views and opinions expressed are those of the author(s) only and do not necessarily reflect those of the European Union or the European Research Executive Agency (REA). Neither the European Union nor the granting authority can be held responsible for them.

ER was supported by an NHMRC Investigator Grant (GNT1195460).

## Declarations of interest

ER has received royalties for co-authoring a chapter on deprescribing in UpToDate. The other authors declare no competing interests.

## Ethics approval

This study was approved by the RCSI University of Medicine and Health Sciences Research Ethics Committee (Reference Number: REC202411011; Dated: 27/01/2025). The study protocol was preregistered on the Open Science Framework (https://doi.org/10.17605/OSF.IO/UTCWB).

## Data availability

All data used in this study are available in the supplementary materials. The analysis scripts are publicly available on Zenodo (Available at: https://zenodo.org/records/21710784).

## Supporting information

FDA Drug class Mapping

EMA Drug class Mapping

Details

Disproportionality analysis based on individual drugs

EMA Disproportionality analysis based on individual drugs

FDA Inter-class disproportionality analysis

EMA Inter-class disproportionality analysis

FDA Intra-class disproportionality analysis

EMA Intra-class disproportionality analysis

FDA Results by age group

EMA Results by age group

FDA Results by sex

EMA Results by sex

EMA Results by country

FDA by timeperiod

EMA by timeperiod

## Notes

### Competing Interest Statement

The authors have declared no competing interest.

### Clinical Protocols

https://doi.org/10.17605/OSF.IO/UTCWB

### Funding

- This research is funded by the European Union through the Marie Skłodowska-Curie Actions (MSCA) programme. Views and opinions expressed are those of the author(s) only and do not necessarily reflect those of the European Union or the European Research Executive Agency (REA). Neither the European Union nor the granting authority can be held responsible for them.: HEAD-P (101149577): https://cordis.europa.eu/project/id/101149577

