## Supplementary material for "Adverse drug withdrawal event signals in FAERS and Eudravigilance databases: a stratified disproportionality analysis study": Details

**Table S1.1:** Relevant and non-relevant Preferred Terms (PTs) containing “withdraw” in databases.

| **Relevant PTs** | **Non-Relevant PTs** |
| --- | --- |
| **PTs in EMA-EV database** | |
| Withdrawal syndrome | Drug withdrawal syndrome neonatal |
| Drug withdrawal syndrome | Alcohol withdrawal syndrome |
| Steroid withdrawal syndrome | Abnormal withdrawal bleeding |
| Drug withdrawal convulsions | Drug withdrawal maintenance therapy |
| Drug withdrawal headache | Thought withdrawal |
| Withdrawal arrhythmia | **-** |
| Withdrawal catatonia | **-** |
| Withdrawal hypertension | **-** |
| **PTs in FDA-FAERS database** | |
| Withdrawal syndrome | Abnormal withdrawal bleeding |
| Drug withdrawal syndrome | Alcohol withdrawal syndrome |
| Drug withdrawal headache | Anti-androgen withdrawal syndrome |
| Drug withdrawal convulsions | Drug withdrawal maintenance therapy |
| Steroid withdrawal syndrome | Drug withdrawal syndrome neonatal |
| Topical steroid withdrawal reaction | Tobacco withdrawal symptoms |
| Withdrawal catatonia | Withdrawal bleed |
| Withdrawal hypertension | Withdrawal of life support |
| Dopamine agonist withdrawal syndrome | Withdrawal Hepatitis |
| - | Thought withdrawal |
| - | Withdrawal bleeding irregular |

**Key variables**

Different key variables were selected based on the study objectives. In FDA FDA-FAERS database, each case is identified by a unique primary identifier (primaryid), whereas in the EMA-EVdatabase, the local report number (evlocalreportnumber) serves as the unique case identifier. Reporting over time were evaluated using the report received date (*fda_dt* in FDA-FAERS and *first_received_date* in EMA-EV). Patient demographic variables included age (*age_grp* in FDA-FAERS; *patient_age_group_id* in EMA-EV) and sex (*sex* in FDA-FAERS; *patient_sex_id* in EMA-EV). ADWEs were identified using the preferred term variable *pt* in FDA-FAERS and *reaction_code–pt* in EMA-EV, both of which capture standardised Medical Dictionary for Regulatory Activities (MedDRA) preferred terms (PTs). Drug information was obtained from the *drugname/prod_ai* variable in FDA-FAERS and *recodedproductsccomps* in EMA-EV. Additionally, the country variable (*country_id*) used only in the EMA-EV database. Detailed descriptions of the key variables from both FDA-FAERS and EMA-EV used in this study are provided in **Table S1.2**.

**Table S1.2:** Details of similar variables available in the FDA-FAERS and EMA-EV databases

| **FDA-FAERS** | | **EMA-EV** | |
| --- | --- | --- | --- |
| **Variable** | **Descriptor** | **Variable** | **Descriptor** |
| **Primaryid** | Unique number for identifying a report | **Evlocalreportnumber** | Unique number assigned to each case |
| **Fda_dt** | Date FDA received case | **First_received_date** | Date on which the report was first received from source |
| **Age** | Numeric value of patient's age at event | **Onset_age** | Age at time of onset of reaction/event (number) |
| **Age_code** | dec = decade;  yr = year;  mon = month;  wk = week;  dy = day;  hr = hour. | **Onset_age_unit** | Age at time of onset of reaction/event (unit) |
| **Age_grp** | n = neonate;  i = infant;  c = child;  t = adolescent  a = adult;  e = elderly | **Patient_age_group_id** | Patient age group |
| **Sex** | m = male;  f = female;  unk = unknown | **Patient_sex_id** | m = male;  f = female;  unk = unknown |
| **Prod_ai** | Product active ingredient (drug). Available during 2014Q3 to 2023Q4 | **Recodedproductsccomps** | Recorded product compound (drug) |
| **drugname** | Valid trade name/Verbatim name as entered by the reporter. Available during 2004Q1 to 2023Q4 | **-** | - |
| **Role_code** | ps = primary suspect drug;  ss = secondary suspect drug;  c = concomitant;  i = interacting | **characterisation_id** | Characterisation of drug role (Suspect, concomitant, and interacting) |
| **PT** | Preferred term for the reaction/event, using the MedDRA | **Reaction_code-pt** | Preferred term for the reaction/event, using the MedDRA |
| **Only EMA** | | **Country_id** | Reporter’s country code |

**Table S1.3:** Data cleaning process (data merging and deduplication)

| **Dataset** | **Primary identifier** | **Deduplication method** |
| --- | --- | --- |
| FDA-FAERS | primaryid | Retain only most recent report based on FDA_DT; remove duplicates using CASE_ID, sex, age, reporter_country, caseversion, and i_f_cod |
| EMA-EV | evlocalreportnumber | Retain only most recent report based on first_received_date; identify duplicates using first_received_date, message_creation_date, onset_age, patient_sex_id, country_id |
| Description of the data cleaning process (data merging and deduplication) | | |
| First, we merged 20-year (2004Q1-2023Q4) FDA (LAERS and FAERS data) and EMA-EV data separately in Stata software. In FDA, we mapped "isr" to "primaryid" (as equivalent entities) when merging the LAERS and FAERS datasets. In the FDA data, each case is identified by a primary identifier (primaryid) whereas the EMA-EV data use a local report number (evlocalreportnumber) as the unique case identifier ***(see Table S1.2)***. To ensure consistency, we retained only the report with the most recent FDA_DT (date on which the case was received/the case received by FDA) for FAERS, and the most recent first_received_date (date on which the report was first received from source/the report is submitted to EV) for EMA-EV. Additionally, the deduplication procedure was performed to identifies reports with identical values across specific fields. The FDA-FAERS dataset required cleaning to identify and remove potential duplicate reports before analysis. This was included CASE_ID (caseid: number for identifying a FAERS case), sex, age, reporter_country (country of the reporter), caseversion (version of the report), and i_f_cod (indicating whether the report is initial or follow-up) for FDA-FAERS.  In EMA, we also conducted deduplication based on first_received_date, message_creation_date (date of message creation), onset_age, patient_sex_id, and country_id to identify duplicate records. This approach ensured that only a single report was retained for each case. Complete details of data cleaning process (merging, cleaning and deduplication) with all Stata commands are available on ***Zenodo (Available at:*** [***https://zenodo.org/records/18155023***](https://zenodo.org/records/18155023)***).*** Files details ***FDA_DataLoop_LAERS_FAERS*** (.do and txt) and ***EMA_DataLoop_EudraVigilance*** (.do and txt) | | |

**Table S1.4:** Contingency table (2×2) and criteria for disproportionality analysis

| **Drug** | **ADWE** | **Not ADWEs** |
| --- | --- | --- |
| Suspected drug of interest | a | b |
| Not a drug of interest | b | d |
| **Methods** | Formulae | Criteria |
| PRR | a / (a + b) / c / (c + d) | Lower bound 95% CI;  Number of reports ≥ 5; PRR≥2 |
| ROR | (a/c)/(b/d) or ad/bc | Lower bound 95% CI;  Number of reports ≥ 5; ROR >1 |
| IC | log_2_ a + 0.5/a^exp^ + 0.5 where a^exp^ = (a + b) ∗ (a + c)/(a + b + c + d) | Lower bound 95% CI;  Number of reports ≥ 5; IC >0 |

*ADWE: Relevant ADWEs PTs; Non-ADWEs: Other ADRs and non-relevant withdrawal PTs; a: N cases with suspected drug of interest and ADWEs; b: N cases with suspected drug of interest but not the ADWEs; c: N cases with ADWEs but not the suspected drug of interest; d: N cases that do not include the ADWEs and the suspected drug of interest; PRR; Proportional reporting ratio; ROR: Reporting odds ratio; CI: Confidence interval; IC: Information component*
